# SPTLC1 p.Leu38Arg, a novel mutation associated with childhood ALS

**DOI:** 10.1101/2022.02.07.22269609

**Authors:** Museer A. Lone, Sen Zeng, Florence Bourquin, Mengli Wang, Shunxiang Huang, Zhiqiang Lin, Beisha Tang, Ruxu Zhang, Thorsten Hornemann

## Abstract

Amyotrophic lateral sclerosis (ALS) is a progressive and fatal neuromuscular disease. Recently, several gain-of-function mutations in *SPTLC1* were associated with juvenile ALS. SPTLC1 encodes for a subunit of the serine-palmitoyltransferase (SPT) - the rate-limiting enzyme in the *de novo* synthesis of sphingolipids (SL).

Here we identified a novel *SPTLC1p*.*L38R* mutation in a young Chinese girl with a signature of juvenile ALS. The patient presented with muscular weakness and atrophy, tongue tremor and fasciculation, breathing problems and positive pyramidal signs. A blood lipid analysis showed overall increased SL levels and particularly a pronounced increase in dihydro-SL species. Increased SL *de novo* synthesis was confirmed in an SPTLC1 deficient HEK cell model that expressed the mutant form. An experimental therapy based on extracts of the fungus *C. cicadae* resulted in a significant lowering of plasma sphingolipids. A subsequent metabolomics analysis identified Myriocin, a potent SPT inhibitor as an active component of the extract. The inhibitory effect of the *C. cicadae* extract on SL *de novo* synthesis was confirmed in a HepG2 cell model. These results suggest that a pharmacological inhibition of SPT could be a therapeutic option for this form of ALS.

## Introduction

Amyotrophic lateral sclerosis (ALS) is a severe neurological disorder with an annual incidence of approximately 1-2.6 cases per 100 000 and a prevalence of approximately 6 cases per 100 000 (1). The disease is characterized by the degeneration of upper (motor cortex) and lower (brainstem/spinal cord) motor neurons resulting in progressive muscle denervation, paralysis, and death. Most forms are sporadic but about 10 percent have a genetic component although with variable penetrance (2). Recently, several mutations in *SPTLC1* have been associated with a juvenile form of ALS (3). *SPTLC1* encodes for a subunit of the enzyme serine-palmitoyltransferase (SPT) that catalyses first and rate-limiting step in the *de novo* synthesis of sphingolipids (SL). Normally, SPT conjugates the substrates palmitoyl-CoA and L-serine in a pyridoxal-5-phosphate (PLP) dependent reaction to form a long chain base (LCB) which is the common structural elements of all SL. The generated LCB is subsequently metabolized to ceramides and finally converted further to complex sphingolipids such as sphingomyelins (SM) and glycosphingolipids (Supplementary Fig. S1).

SPT consist of three core subunits (SPTLC1, 2 and 3) that are dynamically associated with a set of regulatory proteins (ORMDL3, ssSPTa, ssSPTb) that control enzyme activity and substrate specificity (4-6). Under conditions of L-serine deficiency, SPT can also use L-alanine and glycine as alternative substrates, which forms an atypical class of 1-deoxysphingolipids (1-deoxySL) (Supplementary Fig. S1). Several mutations in *SPTLC1* and *SPTLC2* lead to a permanently increased activity with these alternative substrates, resulting in pathologically elevated 1-deoxySL levels that cause the rare Hereditary Sensory Neuropathy type 1 (HSAN1). Clinically, HSAN1 presents with progressive peripheral sensory loss, neuropathic pain and ulcero-mutilations (7-10). In contrast, the recently reported ALS-SPTLC1 mutations are not associated with increased 1-deoxySL formation but instead with an impaired homeostatic control and an overall increased formation of canonical SL (3). Here we report a novel *SPTLC1* mutation found in a Chinese patient, which presented with juvenile ALS and similar changes in the SL profile.

## Materials and methods

### Patient data

Ethical approval for human subject research studies described in this paper was obtained from Institutional Review Board of the Third Xiangya Hospital in Central South University (protocol no. 21096), China. Informed consent from the girl and her parents was obtained for the research and publication of the results of this project.

Physical examination was performed by two neurologists and detailed history developments of diseases were recorded. Nerve electrophysiological examination and Electromyography (EMG) examination were performed. Juvenile ALS was diagnosed according to E1 Escorial criteria (11).

### Genetic analysis

DNA was extracted from venous blood (10 ml) using a standard phenol-chloroform method. Multiplex ligation-dependent probe amplification (MLPA) was used to screen copy number variants and Next Generation Sequencing (NGS) was performed to screen *SPTLC1* mutations. All variants with an allele frequencies > 1% were excluded. Variants were confirmed by Sanger sequencing. Polyphen-2 (http://genetics.bwh.harvard.edu/pph2/), PROVEAN (http://provean.jcvi.org/seq-submit.php/) and SIFT (http://sift.jcvi.org/) were used to predict mutation impact. Phylogenetic conservation was analysed using MutationTaster (http://www.mutationtaster.org). All mutations were validated according to the guidelines of the American College of Medical Genetics and Genomics (ACMG).

#### Lipid analysis

Lipidomics analysis was performed as described earlier (12). The following standards were used for quantification: doxSA(m18:0)(D_3_), SA(d18:0)(D_7_), SO(d18:1)(D_7_), dhCer (d18:0/12:0), Cer(d18:1/12:0), 1-deoxydhCer(m18:0/12:0), 1-deoxyCer(m18:1/12:0), glucCer(d18:1/8:0), SM (d18:1/18:1)(D_9)_, S1P(D_7_). Pooled samples in 5 concentrations were used for quality control. LCB profiling was performed as described here (6).

### *C. cicadae* treatment and patient follow-up

*C. cicadae* extract was prepared in accordance with traditional Chinese medicine (TCM). Three gram of dried *C. cicadae* were powdered, boiled in 200 ml water (20 min) and orally taken in three portions a day. The patient was followed-up biweekly by video calls and monthly visits. Blood samples were obtained at baseline, 4 and 8 weeks. Disease severity was rated according to the Revised Amyotrophic Lateral Sclerosis Functional Rating Scale (ALSFRS-R) (13).

### Cell culture and labelling

HEK293 cells were cultured in Dulbecco’s Medium (DMEM, Sigma-Aldrich, St. Louis, MO, USA) with 10% FCS. Cells were grown at 37°C in a 5 % CO_2_ atmosphere. The generation of HEK293 SPTLC1 knockout cell line is reported earlier(3). Standard molecular biology techniques were used for generation of all plasmid constructs used in the study. Plasmid transfections were performed with lipofectamine 3000 (ThermoFisher Scientific. Transgenic HEK293 cell lines were selected for growth in DMEM media containing 400 µg/ml Geneticin (Thermo Fischer Scientific).

For labelling assays cells were plated at 200,000 cells/ml in six well plates in Dulbecco’s Medium (DMEM, Sigma-Aldrich) with 10% FCS at 37°C in a 5 % CO_2_ atmosphere. At 70% confluence the medium was exchanged with L-serine free DMEM (Genaxxon Bioscience, Ulm, Germany) containing isotope-labelled D_3_-^15^N-L-serine (1 mM) and D_4_-L-alanine (2 mM) (Cambridge Isotope Laboratories, MA, USA). For inhibition studies, *C. cicadae* extract was cleared by centrifugation (1800 RPM, 5 min, RT), aliquoted and dried. The dried pellet was dissolved in 5 ml water/methanol (50/50, v/v) and filtered (0.4 µm) before use. *C. Cicadae* extract was added directly to the media. After 16 hours, the cells were harvested, counted and cell pellets kept frozen at −20°C until analysis.

### Structure modeling

Models were generated by the SWISS-MODEL server using the automated mode and selecting the model with 100 percent coverage and the best global model quality estimate (GMQE) and qualitative model energy analysis (QMEAN) (14-18).

### Statistical analysis

#### Lipid analysis

Data are expressed as mean ± SD. Statistical evaluation was performed using one-way ANOVA with Tukey correction. Values of P<0.05 were considered statistically significant. Statistical analyses were performed with GraphPad Prism 9.0 (GraphPad Software, Inc., San Diego, CA).

### Data Availability Statement

The raw data that support the findings of this study are available from the corresponding authors upon request.

## Results

### Novel SPTLC1p.L38R mutation causes juvenile Amyotrophic Lateral Sclerosis

The patient, a Chinese girl in the first decade of life presented with a juvenile ALS phenotype. She noticed progressive weakness of the lower limbs and frequent falls. She showed mild pes cavus and abnormal steps. She had difficulties in breathing after walking less than 50 meters. The patient showed a waddling gait and atrophy of the small hand muscles as an expression of distal flaccid paresis. Symmetrical planar and thigh muscular atrophy as well as muscle weakness in four limbs and the pelvic girdle muscles was observed. The MRC scores for distal upper and lower limbs were 3/5, and the MRC scores for proximal upper and lower limbs were 4/5 (Table 1). The girl had ankle clonus and hyperreflexia of the upper limbs. Positive Gowers and Trendelenburg signs were indicative for proximal involvement. Fibrillation of the tongue without previously pronounced tongue atrophy indicate a probably more acute than chronic bulbar involvement. Sensation was intact to all modalities. Autonomic nervous system examination was negative. Nerve conduction studies (NCS) revealed a reduced compound muscle action potential (CMAP) of the left median and bilateral peroneal motor nerves. The CMAP in left ulnar and bilateral tibialis motor nerves was normal. Motor nerve conduction velocities (MNCV), sensory nerve action potentials (SNAP) and sensory nerve conduction velocities (SNCV) in the left median, left ulnar, bilateral peroneus and bilateral tibialis motor nerves were normal (Table 1). The EMG showed diffuse ongoing denervation and chronic reinnervation changes in three segments (cervical, thoracic, and lumbar). Pulmonary function was reduced with 56 % forced vital capacity and 60 % forced expiratory volume per second. Next Generation Sequencing and minor allele frequency filtering (<1%) revealed a novel *SPTLC1* missense mutation c.113T>G (p.L38R) (Fig. 1A) that was absent in both parents (Fig. 1A). The p.L38R variant was not reported on gnomAD, 1000genomes database and dbSNP. Residue L38 is highly conserved across species (Fig. 1B) and predicted to be damaging by several in-silico tools.

**Table 1:** Electrophysiological results of the patient.

| Nerve | Median |  | Ulnar |  | Tibial |  | Peroneal |  |
| --- | --- | --- | --- | --- | --- | --- | --- | --- |
|  | L | R | L | R | L | R | L | R |
| MNCV (m/s) | 61.3 | ND | 60.3 | ND | 52.4 | 50.8 | 53.2 | 50.0 |
| CMAP (mV) | 1.2 ↓ | ND | 5.6 | ND | 8.1 | 8.9 | 1.7 ↓ | 1.3 ↓ |
| SNCV (m/s) | 63.6 | ND | ND | 63.8 | ND | ND | 53.3 | 55.2 |
| SNAP (uV) | 45 | ND | ND | 45.0 | ND | ND | 18.0 | 16.0 |

L, left. R, right.
SNAP = sensory nerve action potential (normal range: median nerve $\geq 20.0 \mu\text{V}$ ; ulnar nerve $\geq 17.0 \mu\text{V}$ ; peroneus superficial nerve $\geq 6.0 \mu\text{V}$ ).
SNCV = sensory nerve conduction velocity (normal range: median nerve $\geq 44.0 \text{ m/s}$ ; ulnar nerve $\geq 44.0 \text{ m/s}$ ; peroneus superficial nerve $\geq 41.0 \text{ m/s}$ ).
CMAP = compound muscle action potential (normal range: median nerve $\geq 5.0 \text{ mV}$ ; ulnar nerve $\geq 5.0 \text{ mV}$ ; peroneal nerve $\geq 2.0 \text{ mV}$ ; tibial nerve $\geq 4.8 \text{ mV}$ ).
MNCV = motor nerve conduction velocity (normal range: median nerve $\geq 50.0 \text{ m/s}$ ; ulnar nerve $\geq 50.0 \text{ m/s}$ ; peroneal nerve $\geq 37.0 \text{ m/s}$ ; tibial nerve $\geq 37.0 \text{ m/s}$ ).

**Figure 1:**
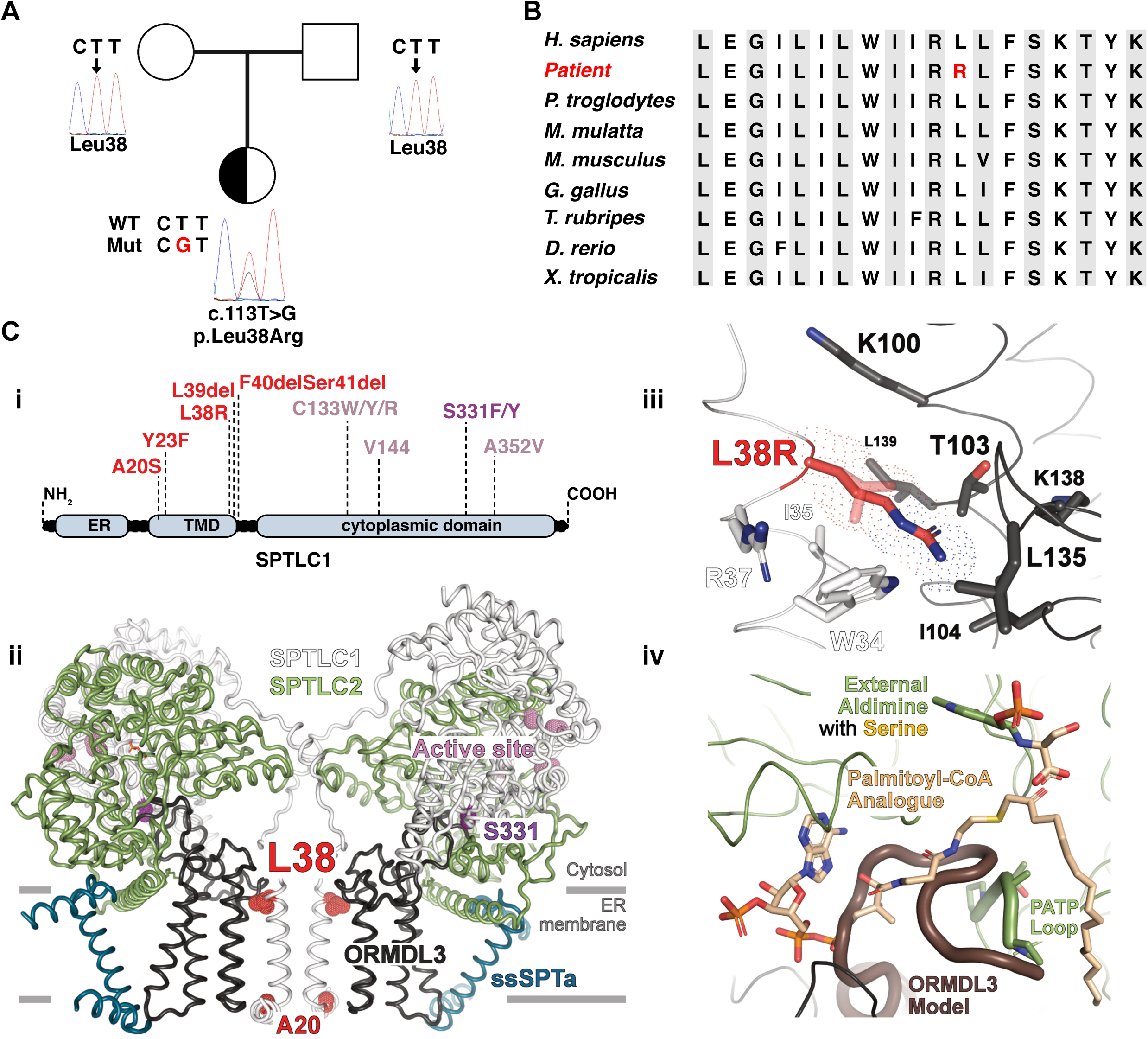
Mutation of the conserved SPTLC1 p.Leu38 causes ALS. (**A**) Pedigree and sequencing chromatograms Square = male; circle = female; half-filled circle = affected child. (**B**) Phylogenetic comparison showed that L38 is highly conserved across species. (**C**) (i) Schematic representation of the SPTLC1-ALS (red) and SPT-HSAN1 (purple) mutations (NM_014941.1). (**C**) (ii) Overall structure of SPT (PDB 7K0M). The purple and red dots represent HSAN1 and ALS mutations, respectively. (**C**) (iii) Close-up on R38 and its environment. The mutation affects the hydrophobic interactions between the transmembrane helices of SPTLC1 and ORMDL3. R38 is 3.7Å longer than L38 and might push the ORMDL3 transmembrane helix away from SPTLC1. R38 is located close to W34 (SPTLC1) and T103 (ORMDL3) as well as the hydrophobic residues I35 (SPTLC1) and L139, L135, I104 (ORMDL3). The mutation may trigger positive charge repulsion and disruption of the Van der Waals interactions between SPTLC1 and ORMDL3. (**C**) (iv) Close-up on the active site, depicting the putative influence of the N-terminus of ORMDL3 on the binding of palmitoyl-CoA and on the conformation of the PATP loop (SPTLC2 499-502). A dissociation of ORMDL3 may alter the position and the conformation of the PATP loop (SPTLC2) to open the palmitoyl-CoA binding site (Fig. 2Div). The full-length ORMDL3 model (dark brown) was generated by the SWISS-MODEL server (https://swissmodel.expasy.org, automated mode and 100% coverage). The global model quality estimate (GMQE) and the qualitative model energy analysis (QMEAN) were 0.81 and −1.77, respectively. All figures were prepared with Pymol (The PyMOL Molecular Graphics System, Version 2.0a0 Schrödinger, LLC).

The p.L38R mutation is located in the N-terminal transmembrane helix motive of SPTLC1 (Fig. 1Ci) that interacts with the regulatory subunit ORMDL3 (Fig. 1Cii). Other mutations within the same region were reported to cause juvenile ALS (3). In contrast, the HSAN1 associated mutations are generally found closer to the active site and PLP binding domain (Fig. 1Ci). To estimate the structural impact of the L38R mutation on the catalytic activity of SPT, the mutation was modelled into the recently published cryoEM structure of SPT (PDB 7K0M) (5). Based on the model, the L38R exchange likely affects the hydrophobic interactions between the transmembrane helices of SPTLC1 and ORMDL3 (Fig. 1Ciii). ORMDL3 is responsible for the homeostatic control, of the cellular SL *de novo* synthesis by regulating the activity of the SPT enzyme A full-length model of ORMDL3 revealed a putative competition of the ORMDL3 N-terminus and the palmitoyl-CoA binding site (Fig. 1Civ). A dissociation of ORMDL3 from SPTLC1 may alter this position and change the conformation of the PATP loop in SPTLC2 to open the palmitoyl-CoA binding site, hindering the hydrophobic interactions in the transmembrane area between SPT and ORMDL3. Consequently, one would expect for the SPTLC1p.L38R variant a reduction or even a complete loss of the homeostatic control by ORMDL3.

### SPTLC1p.L38R mutation increases SL *de novo* synthesis

The effect of the SPTLC1p.L38R mutation on SPT activity was analyzed in the background of an SPTLC1 deficient HEK293 cell line (3). The SPTLC1 knockout (KO) cells were transfected to express either SPTLC1wt, the p.L38R ALS mutant or the p.C133W HSAN1 mutant. Cellular sphingolipid *de novo* synthesis was measured using a stable isotope-based activity assay by quantifying the time dependent incorporation of D_3_, ^15^N-serine (1 mM) and D_4_-alanine (2 mM) in *de novo* formed LCB’s. The incorporation of the labelled amino acid results in stable mass shift of the *de novo* formed SL by +3 Da, which was quantitatively analyzed by LC-MS. KO cells expressing the SPTLC1 p.L38R mutant showed an overall increased SL formation (Fig. 2). Compared to SPTLC1wt expressing cells, the relative changes appeared to be the strongest for saturated dhCer (>15-fold) followed by dhSM (7-fold), Cer (4-fold) and SM (2-fold) (Fig 2A and B). This increase in canonical activity was not seen for the SPTLC1 p.C133W expressing cells. In contrast, the p.C133W expressing cells formed significant amounts of 1-deoxySL, which were not formed in SPTLC1 wild type and p.L38R expressing cells (Fig. 2 C).

**Figure 2:**
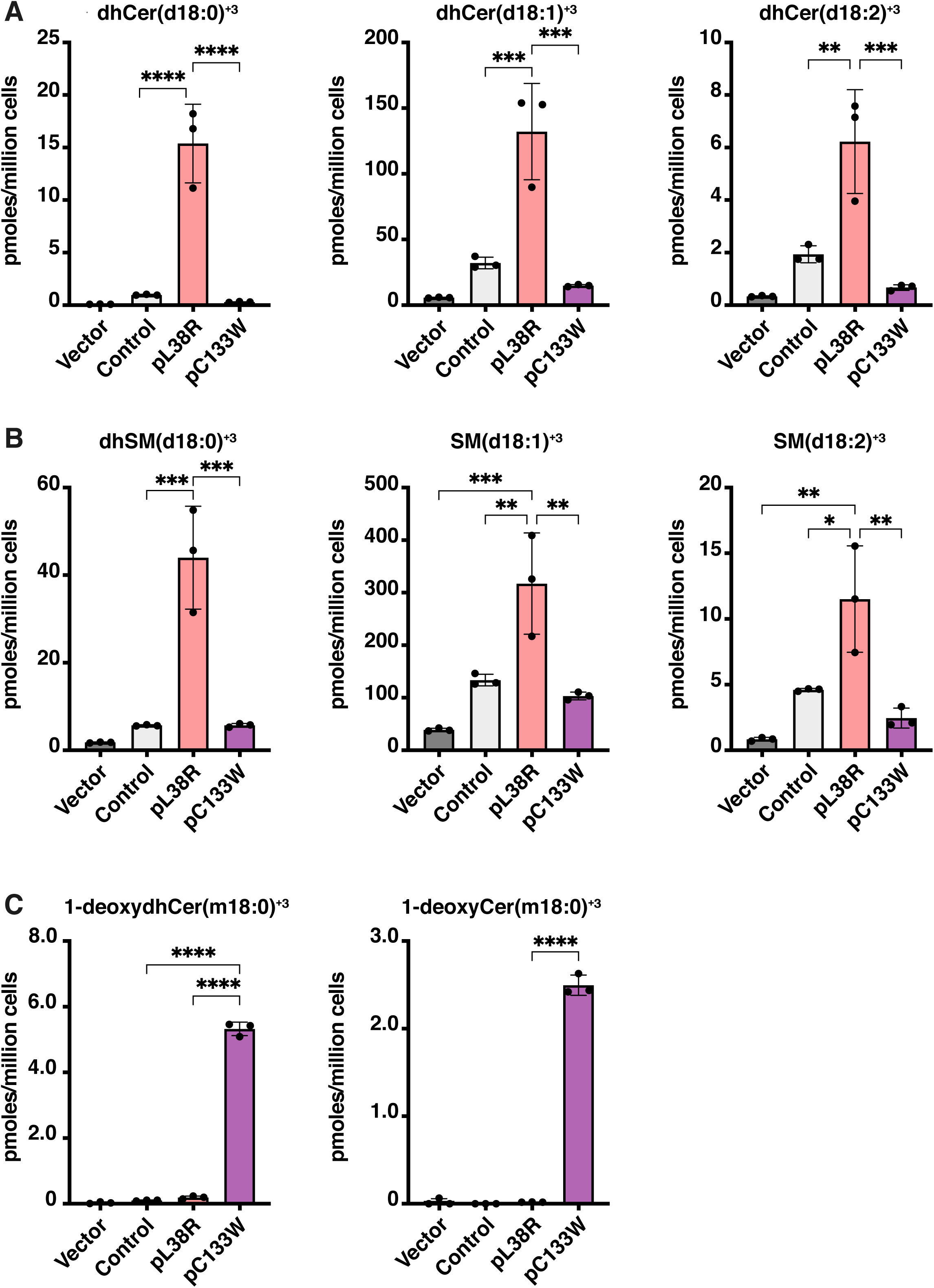
SPTLC1p.Leu38Arg leads to hyperactive SPT enzyme/elevated synthesis of canonical sphingolipid. Expression of the SPTLC1p.L38R mutant in SPTLC1 deficient HEK293 cells. SPT activity was measured by the time dependent incorporation of labelled D_3_, ^15^N-L-serine and D_4_-alanine (16 hours). The SPTLC1p.L38R expressing line showed a significantly increased formation of labelled (**A**) ceramides and (**B**) sphingomyelins whereas (**C**) 1-deoxySLs are only formed in the HSAN1 SPTLC1p.C133W mutant expressing line. Data are shown as mean ± SD. Statistical significance was determined using one way ANOVA followed by Tukey’s correction.

Increased total SL levels were confirmed in patient blood samples. The plasma showed significantly elevated circulating SLs in comparison to the parent, six unrelated HSAN1 patients (SPTLC1p.C133W) and healthy controls (Fig. 3A to D). Again, the changes were most prominent for SL with a saturated LCB (d18:0) than for those containing either one (d18:1) or two (18:2) double bonds (supplementary Fig. S1). Comparing the individual SL classes, the relative increase was more pronounced for Cer (Fig. 3A) and HexCer (Fig. 3B) than for SMs (Fig. 3C). In addition, we also found slightly increased 1-deoxySLs in the patient plasma (Fig. 3D) but in concentrations at the lower end of what is normally seen in HSAN1 (>0.3 µM).

**Figure 3:**
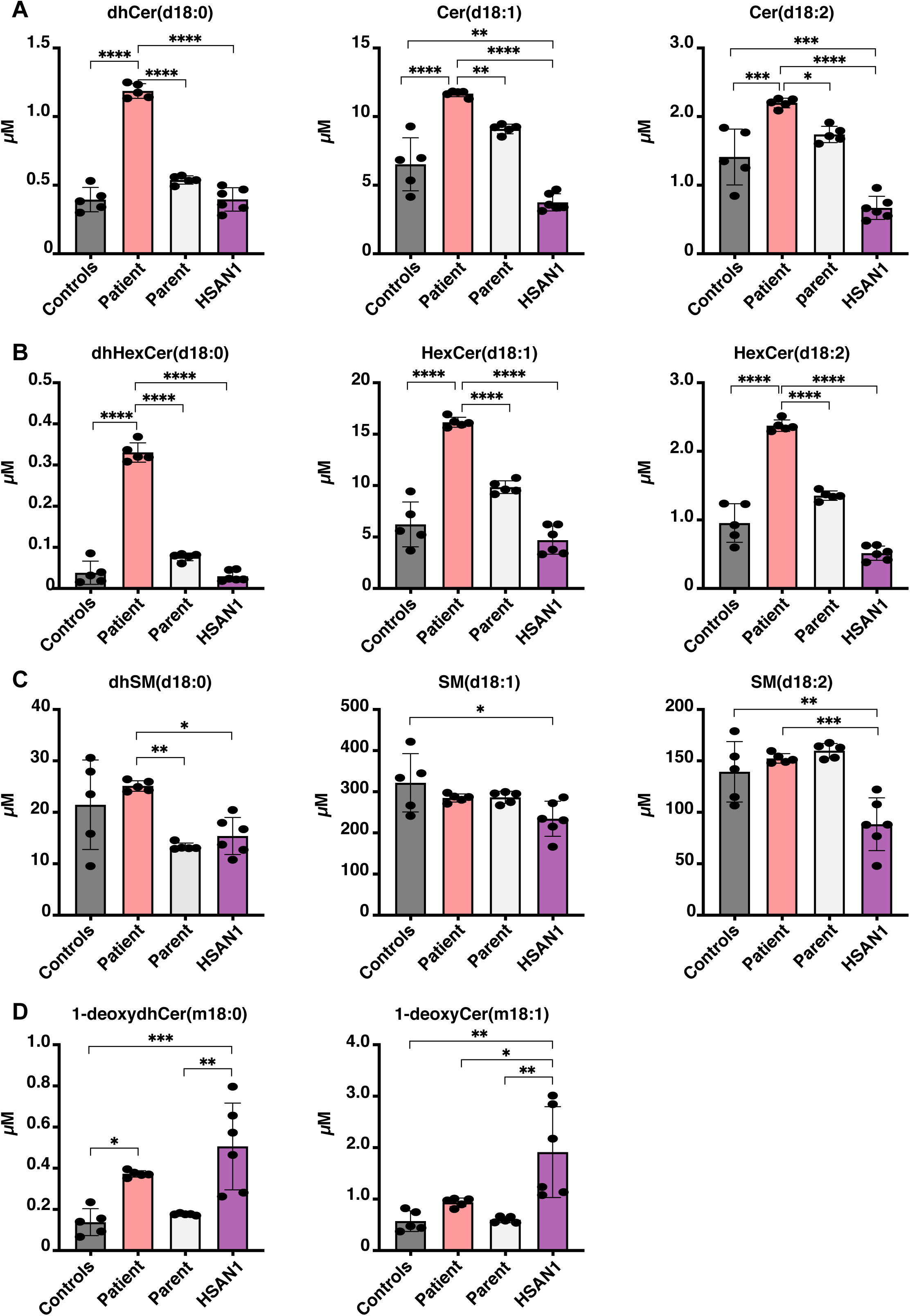
Circulating sphingolipid levels are elevated in SPTLC1p.Leu38Arg patient. (**A**) Ceramide (Cer), (**B**) sphingomyelin (SM), and (**C**) Hexosylceramide (HexCer) levels in the patient relative to her biological parent, a group of unrelated HSAN1 (p.C133W) patients or healthy controls. Mainly Cer and HexCer species with saturated long chain bases (d18:0) were altered followed by mono- (d18:1) and di- (d18:2) saturated species. (**D**) 1-deoxySL are significantly increased in plasma of HSAN1 patients (SPTLC1 p.C133W). Plasma 1-deoxySL in the patient were slightly elevated compared to controls. Data are shown as mean ± SD. Statistical significance was determined using one way ANOVA followed by Tukey’s correction.

### *Cordyceps cicadae* extracts lower circulatory sphingolipid levels in the patient

As there is currently no therapy available for this condition, the responsible neurologists in China decided to treat the patient with an experimental TCM based therapy based on extracts of the entomopathogenic fungi *C. cicadae*. This fungus was reported earlier to form the SPT inhibitor Myriocin (19). As the L38R variant was associated with increased plasma SL levels, we tested whether this approach could reverse this increase. The dried fungi (Fig. 4A) were obtained from a medical TCM provider in Shanxi Provincial Hospital and confirmed as *C. cicadae* by ITS sequencing (Supplementary Fig. 2). The extract was prepared according to the traditional TCM method by boiling in water and applied orally in three portions a day. A metabolomics analysis of the extract using high-resolution MS identified the presence of Myriocin and of other, similar metabolites in the extract (Supplementary Table 1).

**Figure 4:**
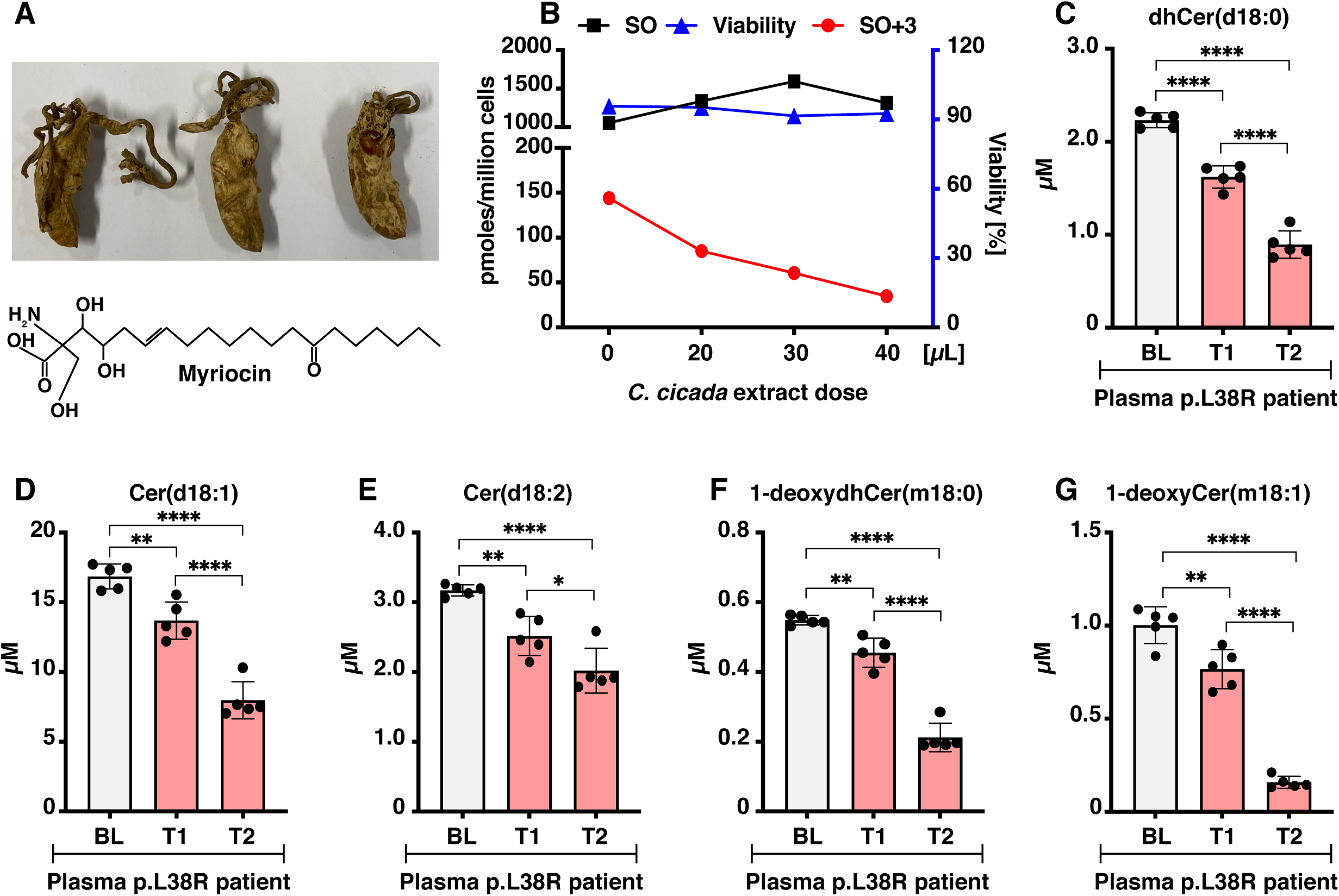
Cordyceps extracts normalize plasma sphingolipid levels in the patient. (**A**) *Cordyceps cicadae* used for preparation of the TCM extract. Chemical structure of SPT inhibitor myriocin (C21H39NO6, 401.3 Da). (**B**) Incorporation of isotope labelled L-serine in long chain base, sphingosine (SO+3) in HepG2 cells is inhibited by the *C. cicadae* extracts in a dose dependent manner. Unlabelled sphingolipids (SO) remain unchanged. (**C** to **F**) The *C. cicadae* extracts decrease sphingolipid levels in the patient girl over time (BL, base line; T1 = 4 weeks after TCM, T2 = 8 weeks after TCM). All data are shown as mean ± SD. Statistical significance was determined using two-way ANOVA with Tukey’s correction.

The inhibitory effect of the extract on SL metabolism was confirmed in a HepG2 hepatocyte model (Fig. 4B). SL *de novo* synthesis was quantified by the incorporation of stable isotope labelled substrates as describe above. Total SPT activity was determined after hydrolyzing the extracted lipids to release the LCB as described previously (6).

We observed a dose dependent reduction of *de novo* formed C_18_SO+3 (Fig 4B) whereas the levels of previously formed unlabelled SO remained unchanged. This indicates that the *C*.*cicadae* extract acts on *de novo* synthesis rather than on the catabolic of the SL metabolism (Fig 4B). The Cordyceps extract had no influence on cell viability (Fig 4B). The plasma SL profile of the patient was analyzed 4 (TP1) and 8 (TP2) weeks after start of the therapy (Fig. 4 and supplementary Fig. S3). We observed a significant and time dependent decrease in plasma ceramide in response to the therapy (Fig. 4C and G). In addition, levels of complex SL were reduced which was more significant for HexCer than for SMs (Supplementary Fig. S4A to F).

## Discussion

The contribution of genetic factors in ALS is still elusive. Recently, several mutations in the SPTLC1 gene of SPT were shown to be associated with early childhood ALS (3). Here, we report a novel SPTLC1p.L38R missense mutation that was identified in a young Chinese girl with juvenile ALS. The mutation is located adjacent to the previously reported SPTLC1p.L39del ALS variant (3). Based on the recently published SPT structure (5), L38 seems to interact directly with the inhibitory subunit ORMDL3 that controls SL homeostasis by regulating SPT enzyme activity. The exchange of an apolar Leu with a polar Arg (Fig. 1A, B and C) alters this interaction. Losing the SPT - ORMDL3 interaction results in a hyperactive SPT enzyme (20). An impaired homeostatic control was confirmed by measuring SL *de novo* synthesis in an SPTLC1 deficient Hek293 cell line that expressed the p.L38R mutant (Fig 2). Here, the most significant changes were seen for dhCer (d18:0) species followed by Ceramide with either one (d18:1) and two (d18:2) double bonds (Fig 2A and B). This order reflects the metabolic conversion of SLs, starting with the formation of a saturated LCB followed by the introduction of first a Δ4E (by DEGS1) and a second a Δ14Z (by FADS3) double bond (Supplementary Fig. 1).

Blood samples of the patient showed elevated SL levels when compared to either the non-affected parent or European controls. Like in the cell model, mainly Cer and HexCer species with saturated long chain bases (d18:0) were altered followed by mono- (d18:1) and di- (d18:2) saturated species (Fig. 3A to D). SL with a saturated LCB are generally minor but elevated levels have been reported in ORMDL3 deficient mice (20) and in context of a rare form of leukodystrophy that is caused by Ceramide Desaturase 1 (DEGS1) deficiency (21).

It is surprising that a systemic mutation in a ubiquitously expressed enzyme such as SPT can specifically cause a motor neuron disease without other more systemic defects. In addition to the increased canonical SLs we also detected slightly elevated 1-deoxySLs the plasma (Fig. 3D). 1-deoxySLs are neurotoxic and associated with peripheral sensory loss (HSAN1). However, the observed increase was minor in comparison to the changes seen for canonical SL and the patient did not show from sensory symptoms. Increased 1-deoxySL formation was also not seen in the mutant overexpressing HEK cell model, indicating that the SPTLC1-L38R mutation is not per se causing a significant shift in the substrate affinity of the enzyme.

As there was no established therapeutic option available, the responsible physicians in China decided to treat the patient with an experimental TCM based approach, based on aqueous extracts of the fungi *C. cicada*. Cordyceps based extracts are traditionally used in East Asian for health food and herbal medicines and have been applied in humans for more than a millennium (22). In TCM, *C. cicadae* is used against infections and several chronic diseases (22, 23). Interestingly, certain species of Cordyceps sp. contain Myriocin, a highly specific SPT inhibitor that blocks enzyme activity already at very low concentration (Ki ∼ 0.28 nM). However, the therapeutic use of Myriocin as a pure compound is restricted due to its unfavorable pharmacokinetic properties and severe side effects (24). In contrast, TCM based *C. cicadae* extract is generally tolerated well and not known to cause significant adverse effects (22, 23). Rodent studies illustrate that Cordyceps extracts are safe for mammals (25). An untargeted metabolomics analysis confirmed the presence of Myriocin but also of other potential Myriocin derivatives in the extract (Supplementary Table 1). This suggests that maybe other bioactive metabolites in the extract could also contribute to the effect on SL metabolism (26).

In response to the TCM treatment, we observed a significant reduction in circulating SL in the plasma of the patient. Reciprocal to their formation, saturated SL were reduced most followed by SL with mono- (d18:1) and di- (d18:2) unsaturated backbones (Fig. 4C and supplementary Fig S3A and D). The inhibitory effect of the extract on SL *de novo* synthesis was confirmed in HepG2 hepatocytes, which showed a dose dependent reduction in SPT activity and SL *de novo* synthesis (Fig. 4B).

In conclusion, our results support the finding that juvenile ALS can be caused by a hyperactive SPT resulting in a non-properly controlled SL homeostasis. In addition, we showed that a limited pharmacological inhibition of the hyperactive enzyme could be a therapeutic approach for this form of ALS.

## Data Availability

All data produced in the present work are contained in the manuscript.

## Acknowledgments

We are grateful to the patient and her families.

## Author contributions

Conceptualization: MAL, SZ, RZ, TH

Methodology: MAL, SZ, MW, BT, FB

Patient follow up: SZ, ZL

Nerve electrophysiologic and EMG examination: SH

Funding acquisition: MAL, RZ, TH

Writing – original draft: SZ, MAL, RZ, TH

Writing – review & editing: SZ, MAL, RZ, TH

## Supplementary Materials

Fig. S1 to S3

Tables S1

