## Supplementory Fig. S1, Supplementory Fig. S2, Supplementory Fig. S3, Supplementory Table S1 for "SPTLC1 p.Leu38Arg, a novel mutation associated with childhood ALS"

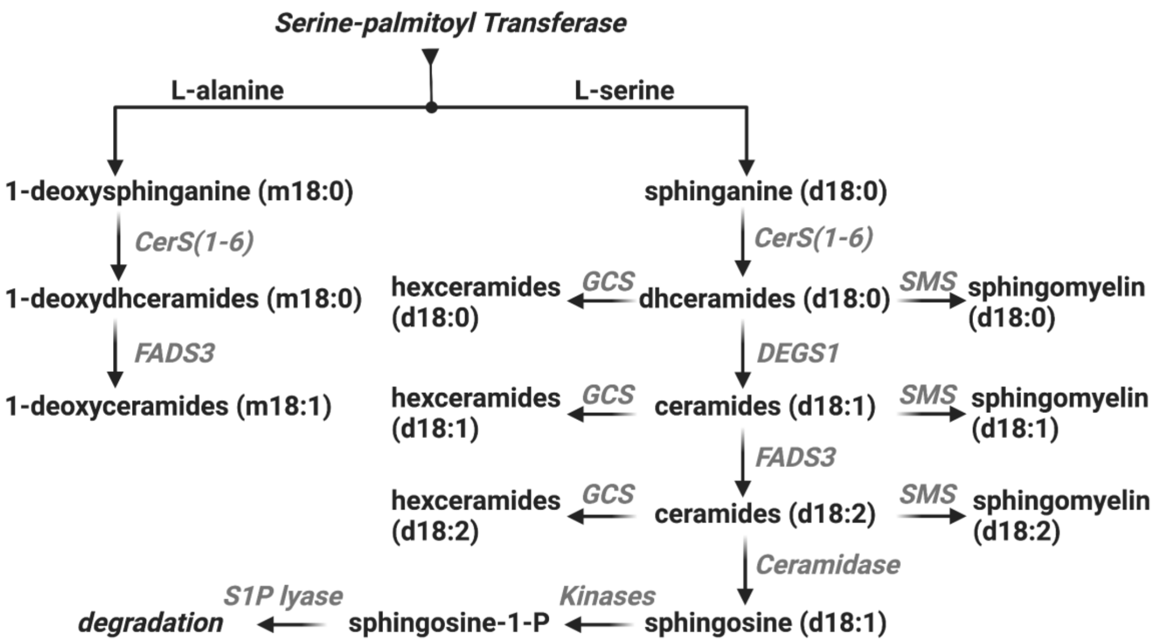


**Figure S1: The *de novo* synthesis of canonical and 1-deoxysphingolipids.**

The serine palmitoyltransferase **(**SPT) catalyzes the first and rate-limiting reaction in the *de novo* synthesis of sphingolipids. It typically conjugates palmitoyl-CoA and L-serine forming sphinganine (SA, d18:0). SA is subsequently N-acylated by ceramide synthases (CerS1-6) forming dihydroceramides (dhCer, d18:0). A Δ4E double bond is introduced by the dihydroceramide desaturase (DEGS1) forming ceramides (Cer, d18:1). Partly, a second Δ14Z double bond is introduced by the fatty acid desaturase 3 (FADS3) forming d18:2 ceramides (Cer, d18:2). Saturated and non-saturated ceramides can be converted to complex sphingolipids, including sphingomyelin (SM) and hexosylceramides (HexCer). In its alternative reaction, SPT can also metabolize L-alanine forming 1-deoxyceramides that are not converted to complex sphingolipids.

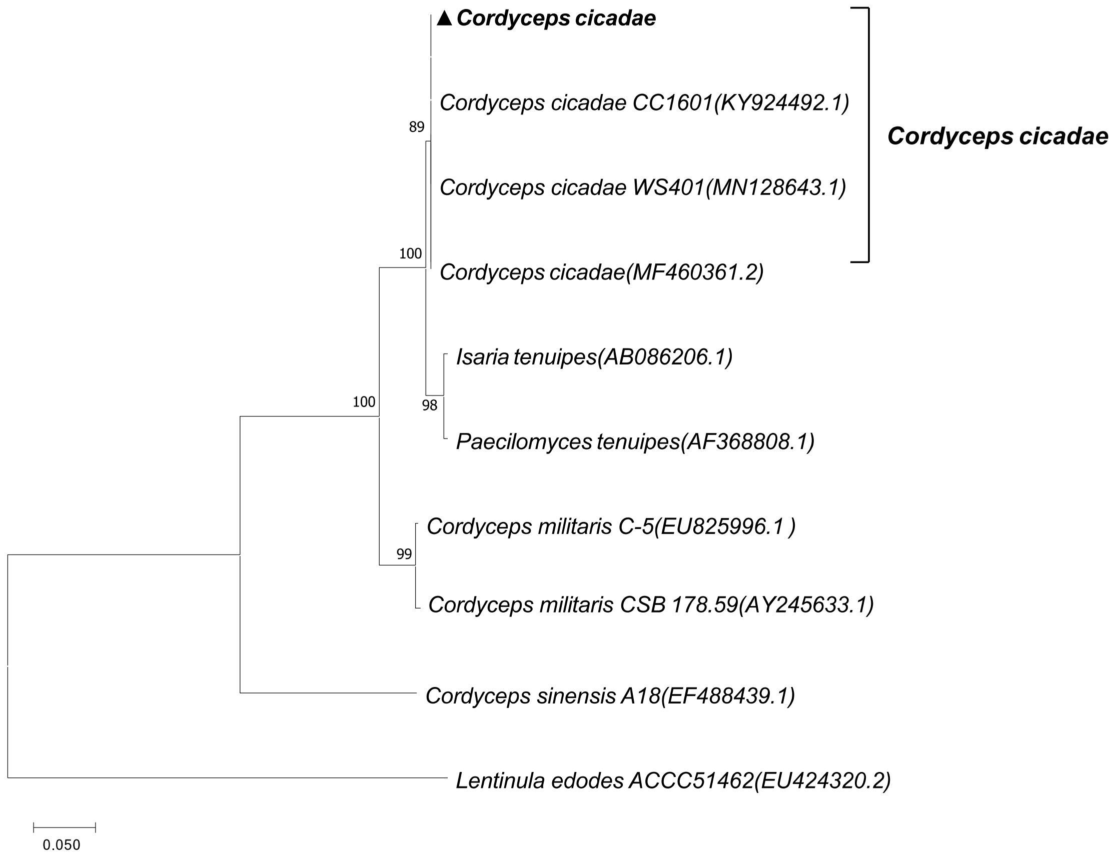


**Figure S2: Phylogenetic tree of fungi from NJ analysis of the ITS sequences using MEGA.** In order to classify the sub-strain of the fungi strain. We extracted genomic DNA from the fungi strain, performed sequencing of ITS sequence, performed sequence alignment with NCBI-Blast and constructed the phylogenetic tree. The reference sequences are retrieved from NCBI-Genbank and the evolutionary history was inferred using Neighbor-Joining method in MEGA version X. The evolutionary distances were computed using the Kimura 2-parameter method. The results show that the fungi completely share the ITS sequence with several kind of *Cordyceps cicadae* strains and the fungi is located to the same phylogenetic branch with *Cordyceps cicadae*.

**Table S1: Myriocin and myriocin related compounds detected in the *Cordyceps cicadae* extracts by metabolomics**

| Molecular formula | Matching compound | Calculated molecular weight | Annot. DeltaMass [ppm] | RT [min] |
|---|---|---|---|---|
| C21 H39 N O5 | Mycestericin D or E | 385.28303 | 0.55 | 12.329 |
| C21 H39 N O6 | Myriocin | 401.27736 | -0.93 | 13.504 |
| C21 H41 N O5 | Mycestericin F or G | 387.29829 | -0.48 | 5.224 |
| C21 H41 N O6 | Mycestericin B or C | 403.29366 | 0.67 | 11.046 |
| C21 H41 N O6 | Mycestericin B or C | 403.29362 | 0.58 | 9.237 |
| C21 H41 N O6 | Mycestericin B or C | 403.29461 | 3.02 | 11.158 |
| C21 H41 N O6 | Mycestericin B or C | 403.29378 | 0.97 | 8.057 |

The following software and databases were used: Compound Discoverer 3.2 (Thermofischer) MSDIAL-insilicoMSMS-Lipids (pos/neg) (2019), Metlin mass spectral database (Scripps Center for Metabolomics) after acquiring the spectra on a Q-Exactive MS analyzer (Thermo Scientific)

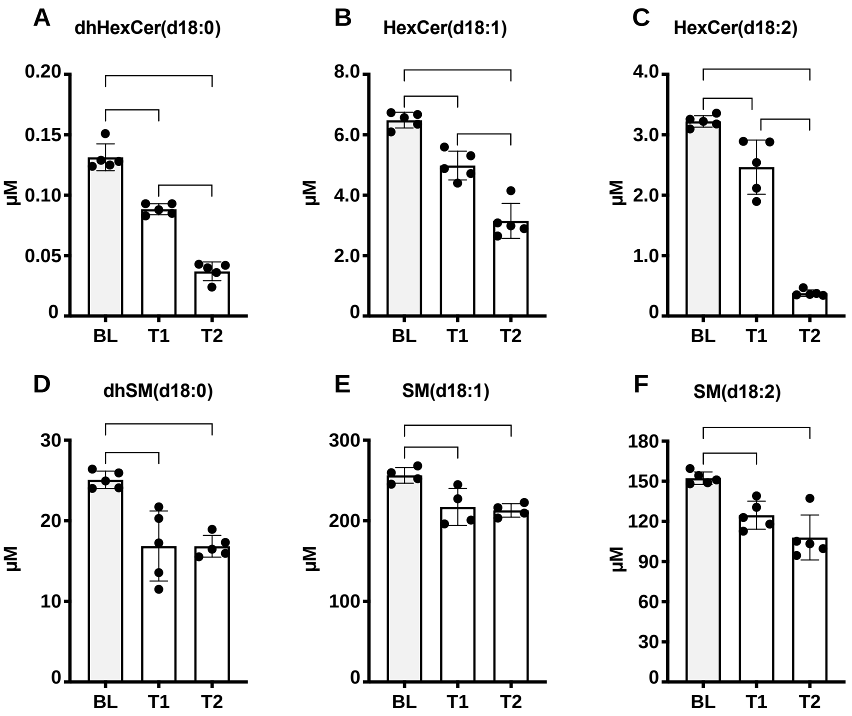


**Figure S3: Effect of Cordyceps extract on complex sphingolipids in plasma.** The *C. cicadae* extracts decrease levels of complex sphingolipids, (A to C) Hexosylceramides (HexCer) and (D to E) Sphingomyelins (SM) in the patient girl over time (BL, base line; T1 = 4 weeks after TCM, T2 = 8 weeks after TCM). All data are shown as mean ± SD. Statistical significance was determined using two-way ANOVA with Tukey's correction.
